# Barriers and Enablers to Physical Activity in Adults and Children with Type 1 Diabetes: A National Survey and Theoretical Mapping

**DOI:** 10.1101/2025.08.05.25333022

**Authors:** Emma. J. Cockcroft, Chris Bright, Hilary Nathan, Dan Farrow, Catherine Russon, Richard Pulsford, Robert. C. Andrews

## Abstract

**Objectives:** To identify and compare barriers and enablers influencing physical activity (PA) in adults and children with type 1 diabetes (T1D) in the UK, and to map these factors to behavioural theory to inform intervention development.

**Methods:** A cross-sectional, national online survey was distributed via Breakthrough T1D (formerly JDRF) networks between December 2022 and January 2023. The survey included closed and open-ended questions on PA behaviours and influencing factors. Responses were collected from adults with T1D and parents of children with T1D. Quantitative data were analysed using descriptive statistics and two-proportion z-tests. Free-text responses were analysed thematically. Barriers and enablers were synthesised using the COM-B model and socioecological framework to identify behavioural determinants and intervention targets.

**Results:** 311 responses were analysed (182 adults, 129 parent proxies for children). Stigma and negative comments were reported by nearly three quarters of both groups. Adults were more likely to cite clinical and motivational barriers, notably fear of hypoglycaemia (53%) and lack of motivation (39%). Children more frequently reported external barriers such as lack of education among coaches and organisers (31%). Motivators differed by age, with adults prioritising health and diabetes management, and children emphasising enjoyment and social aspects. Mapping to COM-B and TDF highlighted the need for interventions addressing both personal and environmental factors.

**Conclusion:** Findings highlight the need for multi-level interventions to address stigma and stereotyping. Educational initiatives and policy measures are essential to improve understanding of T1D, ensure reasonable adjustments, and promote equity and inclusion across settings.

**What is already known?:**

- People with type 1 diabetes (T1D) face unique barriers to physical activity, including fear of hypoglycaemia and the complexity of diabetes management.
- Most existing studies focus on either adults or children and overlook social and environmental factors.

**What this study adds?:**

- This is the first UK-wide study to compare barriers and enablers to physical activity in adults and children with T1D.
- This survey identifies distinct age-related challenges and highlights stigma and lack of support in sport settings as major, under-recognised barriers.

**How this study might affect research, practice or policy:**

- Findings support the need for multi-level interventions and policy reform to address stigma and improve diabetes education among coaches, teachers, and healthcare professionals.

## 1. INTRODUCTION

Type 1 diabetes (T1D) is a chronic autoimmune condition that affects 9.5 million individuals globally (1), with significant prevalence among both children and adults (2). Management of T1D hinges on a complex interplay of insulin administration, diet, lifestyle and blood glucose monitoring. Among these strategies, physical activity (PA) (such as walking, cycling, or playing sport) is a cornerstone, not only for maintaining glycaemic control but also for reducing long-term cardiovascular risks and enhancing mental well-being (3,4). Despite these well-documented benefits, people with T1D often face considerable barriers to achieving recommended PA levels (5,6).

Research consistently highlights that individuals with T1D engage in less PA compared to their peers without T1D. For children and adolescents with T1D, less than half meet the recommended 60 minutes of moderate-to-vigorous PA (MVPA) daily (7). Adults with T1D also report lower levels of engagement in structured exercise and recreational activities (8), with only approximately one third meeting PA recommendations (9), of 150 min or more of MVPA per week (10). This disparity underscores the existence of unique challenges tied to T1D management during PA, such as fears of hypoglycaemia, the need for precise insulin adjustments, and logistical concerns related to monitoring blood glucose levels (6,11,12). This is in addition to commonly cited barriers to PA at a whole population level such as environmental factors, time, and social norms (13).

Theoretical models provide a structured lens for understanding and changing health-related behaviours. The Capability, Opportunity, and Motivation model of Behaviour (COM-B) conceptualises behaviour as the result of interactions between an individual’s capability, opportunity, and motivation (14). Detailing the factors that influence behaviour within these three domains enables researchers and practitioners to identify modifiable targets for intervention and design strategies that are more likely to be effective. However, behaviour does not occur in isolation. The socioecological model (15) complements COM-B by recognising that individual behaviours are shaped by multiple, interacting layers including interpersonal relationships, organisational settings, community norms, and broader policy environments. Integrating the socioecological perspective with COM-B allows for a more comprehensive understanding of the barriers and enablers to PA in people with T1D, highlighting not only personal capabilities and motivations but also the critical role of social, environmental, and systemic factors.

Despite increased awareness of the challenges relating to PA in T1D, there remains limited understanding of how barriers and motivators interact across different groups and settings. Existing evidence is often fragmented with many studies focused on either adults or children, making it difficult to compare experiences across the lifespan. While some large quantitative studies exist, many have relied on tools such as the Barriers to PA in Type 1 Diabetes (BAPAD-1) questionnaire (16,17), which restricts responses to predefined barriers and may miss important or context-specific factors. Additionally, social and environmental barriers—such as stigma, lack of support, and inadequate awareness in schools and sports settings remain underexplored, particularly at scale.

This study addresses these gaps through a large national survey exploring the lived experiences of adults and children with T1D in the UK. It is the first to directly compare perceived barriers and enablers across age groups, using open-ended questions to capture a broad range of individual, social, and environmental influences. This approach enhances generalisability and highlights novel themes that are often missed in checklist-based surveys, offering valuable insights to inform targeted, multi-level interventions.

## 2. METHODS

This study presents a secondary analysis of anonymised survey data exploring PA experiences in people with T1D. The survey was developed and hosted by NFP research (https://nfpresearch.com) in partnership with BreakthroughT1D UK (formerly Juvenile Diabetes Research Foundation (JDRF)), the UK arm of the global charity BreakthroughT1D, which is dedicated to curing type 1 diabetes and improving the lives of those affected by the condition. No new data were collected; analysis remained within the originally agreed scope. According to institutional and national guidelines, secondary analysis of anonymised data did not require further ethical approval or consent. The research adhered to the Declaration of Helsinki and institutional policies.

Participants rated statements about PA using a five-point Likert scale, with additional multiple-choice and free-text items capturing motivations, barriers, stigma experiences, and information sources. The full survey is provided in the supplementary materials.

### Sample

Survey data were collected from Dec 2022 to Jan 2023. The survey was distributed by Breakthrough T1D (formally JDRF) via their network, inclusive of social media. The participants were a convenience sample of willing participants. All people in the UK with T1D were eligible to participate. Because of the nature of recruitment efforts, it is not possible to determine the total number of potential participants approached to complete this survey.

### Data analysis

Survey responses were summarised using descriptive statistics. Continuous variables are presented as means (± SD), and categorical variables as percentages. Analyses were conducted in Python 3.9 (JupyterLab v3, Anaconda 2023), with two-proportion z-tests used to compare adults and children. Multiple comparisons were addressed using the Benjamini–Hochberg false discovery rate procedure.

Free-text responses were analysed thematically in NVivo (v12), following Braun and Clarke’s approach (14). One researcher initially coded the responses, grouping similar codes into broader categories, which were refined in discussion with the wider team.

Quantitative and qualitative findings were synthesised to identify intervention targets, guided by the COM-B model and socioecological framework. This approach enabled a nuanced understanding of key behavioural determinants for adults and children, distinguishing between individual and external factors.

### Patient and public involvement

This study was co-designed and delivered in collaboration with BreakthroughT1D UK, a national diabetes charity. People with lived experience of type 1 diabetes were involved throughout the project, including shaping survey content, interpreting findings, and identifying practical implications. The research team includes individuals with direct personal experience of T1D, ensuring that analysis and recommendations were grounded in real-world relevance and priorities.

## 3 RESULTS

### Sample characeristics

A total of 311 people responded to the survey, including 182 adults and 129 parent-proxy responses for children (<18 years). Respondents were geographically distributed across all UK nations and regions, with most identifying as White British (82%). The sample was 47% male, 52% female, and <1% non-binary. Use of diabetes technology was common, with 60% using CGM and 53% using insulin pumps. Age was approximately normally distributed (skew = 0.33), with a mean (SD) age of 30.3 (18.2) years. Full demographic characteristics, including disease duration, gender, technology use, ethnicity, and location, are summarised in Table 1.

**Table 1.** characteristics of survey respondents

|  |  | Total sample | Adults | Children |
| --- | --- | --- | --- | --- |
| **Total (n, %)** |  | 311 | 182 (59%) | 129 (41%) |
| **Age (Mean, SD)** |  | 30.3 ± 18.2 | 42.9 ± 12.5 | 12.4 ± 5.0 |
| **Years since diagnosis** |  | 11.2 ± 9.4 | 16.0 ± 9.1 | 4.4 ± 4.3 |
| **Gender** | Male | 146 (47%) | 71 (39%) | 75 (58%) |
|  | Female | 161 (52%) | 110 (60%) | 51 (39%) |
|  | Non-binary | 2 (<1%) | 1 (1%) | 1 (1%) |
|  | Not reported | 2 (<1%) | - | 2 (2%) |
| **Use of technology** | Pump | 164 (53%) | 90 (49%) | 74 (57%) |
|  | Closed loop | 75 (24%) | 47 (26%) | 28 (22%) |
|  | CGM | 186 (60%) | 89 (49%) | 97 (75%) |
| **Ethnicity (% white British)** |  | 254 (82%) | 152 (84%) | 102 (79%) |
| **Geographical location** | East Midlands | 13 (4.1%) | 7 (3.8%) | 6 (4.7%) |
|  | East of England | 20 (6.3%) | 10 (5.5%) | 10 (7.8%) |
|  | London | 36 (11.4%) | 25 (13.7%) | 11 (8.5%) |
|  | North East | 15 (4.8%) | 11 (6.0%) | 4 (3.1%) |
|  | North West | 30 (9.5%) | 19 (10.4%) | 11 (8.5%) |
|  | Northern Ireland | 9 (2.9%) | 4 (2.2%) | 5 (3.9%) |
|  | Scotland | 33 (10.5%) | 28 (15.4%) | 5 (3.9%) |
|  | South East | 64 (20.3%) | 34 (18.7%) | 30 (23.3%) |
|  | South West | 28 (8.9%) | 12 (6.6%) | 16 (12.4%) |
|  | Wales | 11 (3.5%) | 5 (2.7%) | 6 (4.7%) |
|  | West Midlands | 16 (5.1%) | 7 (3.8%) | 9 (7.0%) |
|  | Yorkshire/Humberside | 17 (5.4%) | 6 (3.3%) | 11 (8.5%) |
|  | Other | 15 (4.8%) | 111 (6.0%) | 4 (3.1%) |

Participants reported taking part in a variety of activities, with football, walking and swimming being the most common in children and walking, running and gym attendance being the most cited in adults. Data suggest that individual / self-paced activities were most often reported in adult respondents (15 of top 20 reported activities) than in children (6 of the top 20 reported activities).

### Attitudes, knowledge and self-efficacy for physical activity

Adults were more likely to want to engage in more PA with 65% of adults strongly agreeing or agreeing to the statement “I would like to engage in more PA” compared to 42% of children. Both adults and children (98% and 91% respectively) either strongly agreed or agreed that it is important for people living with T1D to engage in PA.

Motivations for PA varied significantly between adults and children (see figure 1). While health and well-being were the most commonly reported drivers overall, adults were significantly more likely than children to be motivated by general health (78.0% vs. 58.9%, p < .001, adjusted p = .001), mental health benefits (69.2% vs. 51.2%, p = .001, adjusted p = .002), glucose management (46.2% vs. 25.6%, p < .001, adjusted p = .001), body image concerns (36.3% vs. 17.8%, p < .001, adjusted p = .001), daily-life needs (18.1% vs. 6.2%, p = .002, adjusted p = .004), financial or career goals (8.5% vs. 1.6%, p = .004, adjusted p = .006), and feelings of guilt (“I should be active”) (16.5% vs. 6.2%, p = .006, adjusted p = .009).

**Figure 1.**
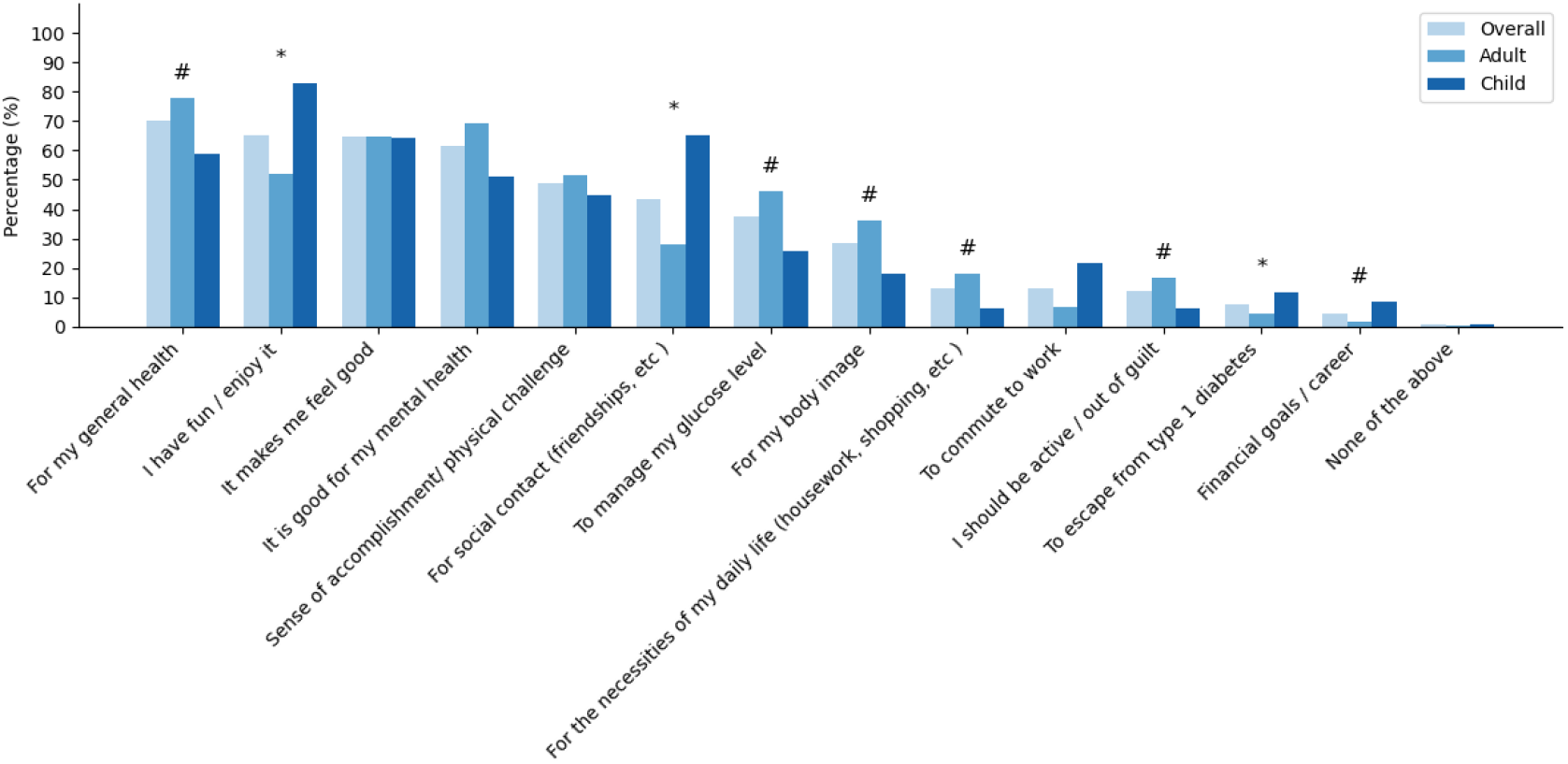
Reasons for engaging in physical activity. Significant differences between adults and children are shown with a # where adults were more likely to report motivation reason and ^*^ where children were more likely to report reason.

Children were significantly more likely to be motivated by fun and enjoyment (82.9% vs. 52.2%, p < .001, adjusted p < .001), social contact (65.1% vs. 28.0%, p < .001, adjusted p < .001), and escaping from type 1 diabetes (11.6% vs. 4.4%, p = .016, adjusted p = .021) compared to adults.

In terms of knowledge and understanding, 30% of adults and 34% of children felt they did not know enough about PA, and 42% and 38% of adults and children respectively would like to be more informed about how they could engage in PA. Additionally, 63% of adults and 65% of children felt that the knowledge of T1D in those organising exercise is not sufficient enough to support them.

### Barriers to engaging in physical activity

Barriers to PA reported by participants are shown in figure 2 (individual level factors) and figure 3 (external factors). The most frequently endorsed barrier was fear of hypo-/hyperglycaemia (46.0%), followed by complexity of diabetes management (39.2%), insufficient time to exercise (33.4%), and lack of motivation (30.9%). Relating to external factors, approximately one-fifth of respondents indicated a lack of education or awareness among coaches and organisers (21.9%), and 16.1% cited insufficient support in their physical-activity settings. Less common barriers included cost (12.2%), never having built an exercise habit (10.6%), and not enjoying exercise (8.7%); stigma or stereotyping was reported as a barrier to PA by 8.0%, and negative messaging from healthcare professionals by only 1.6%.

**Figure 2.**
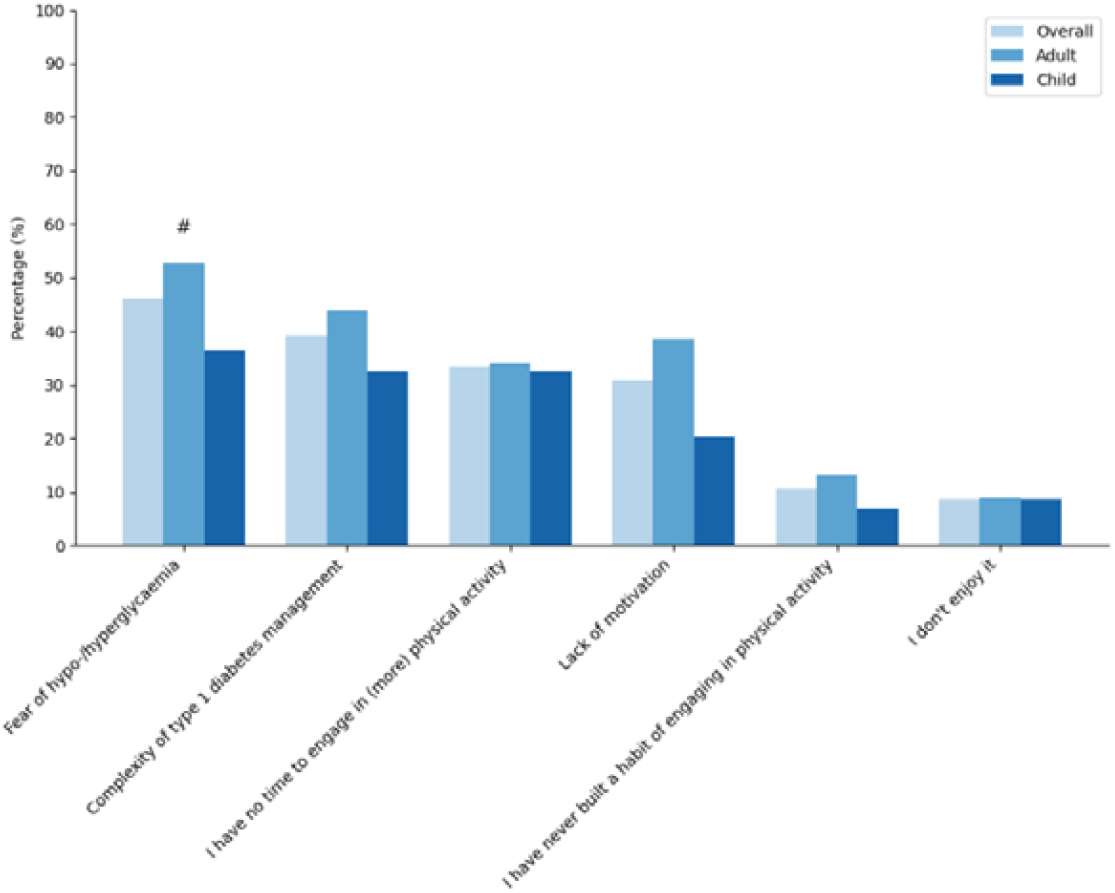
Cited individual level factors for not engaging in more physical activity. Factors cited as barriers for PA at the individual level. Significant differences between adults and children are shown with a # where adults were more likely to report motivation reason and ^*^ where children were more likely to report reason.

**Figure 3.**
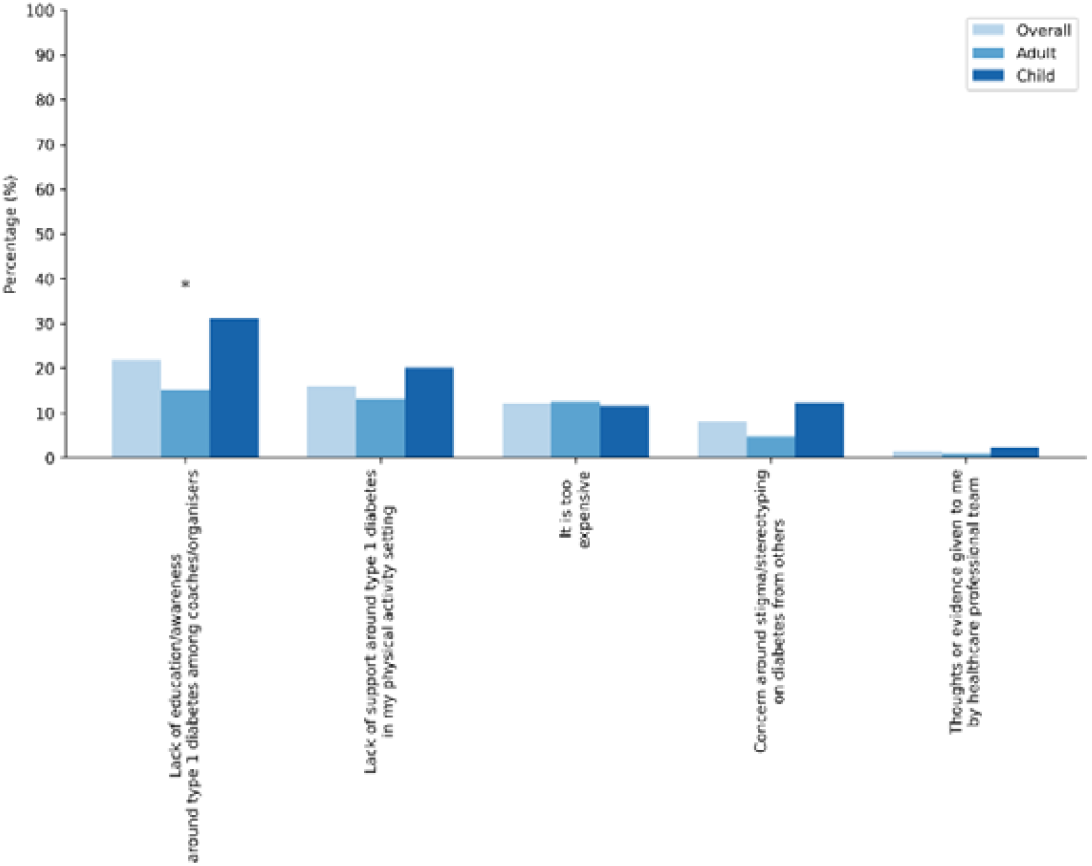
Cited External factors for not engaging in more physical activity relating to internal factors. External barriers to PA .Significant differences between adults and children are shown with a # where adults were more likely to report motivation reason and ^*^ where children were more likely to report reason.

When comparing adults and children, three barriers differed significantly. Adults were more likely to report fear of hypo-/hyperglycaemia (52.7% vs. 36.4%, p = .004, *adjusted* p-value= .018) and lack of motivation (38.5% vs. 20.2%, p < .001, adjusted p-value= .006), whereas children were more likely to report a lack of education or awareness among coaches and organisers (31.0% vs. 15.4%, p = .001, *adjusted* p-value = .006). These results suggest that adults are chiefly impeded by physiological and motivational challenges, while children face additional obstacles stemming from inadequate coach education and support.

### External experiences influencing physical activity

A substantial proportion of participants reported experiencing stigma related to type 1 diabetes and PA.

Overall 73% of participants reported experiencing other people being ignorant about T1D at some point. This was reported to happen ‘regularly’ or ‘very often’ by 27% of participants (21% of adults and 36% of children). Approximately 50% of respondents had heard comments relating to people living with T1D not being able to be physically active, this was reported to happen regularly and very often in 7% of participants (5% of adults, 10% children). Similarly 58% of participants (51% and 68% for adults and children respectively) have experienced coaches, PE teachers or gym instructors expecting less of them because of their T1D. This was reported to happen regularly or very often in 6% of respondents with higher rates in children compared to adults (2% adults vs. 12% children). 47% of participants (50% adults, 41% children) have had people comment on body image in relation to T1D. This was reported to happen regularly or very often in 10% of both adults and children. 73% of participants have had others comment on their diabetes technology in relation to PA (73% of both adults and children). This was reported in 20% of participants (18% of adults and 23% of children). Over half of the participants (48% and 59% for adults and children respectively) had received comments about T1D being their own fault. This was common in 12% of participants (8% of adults and 18% of children). Further breakdown of results shown in figure 4.

**Figure 4.**
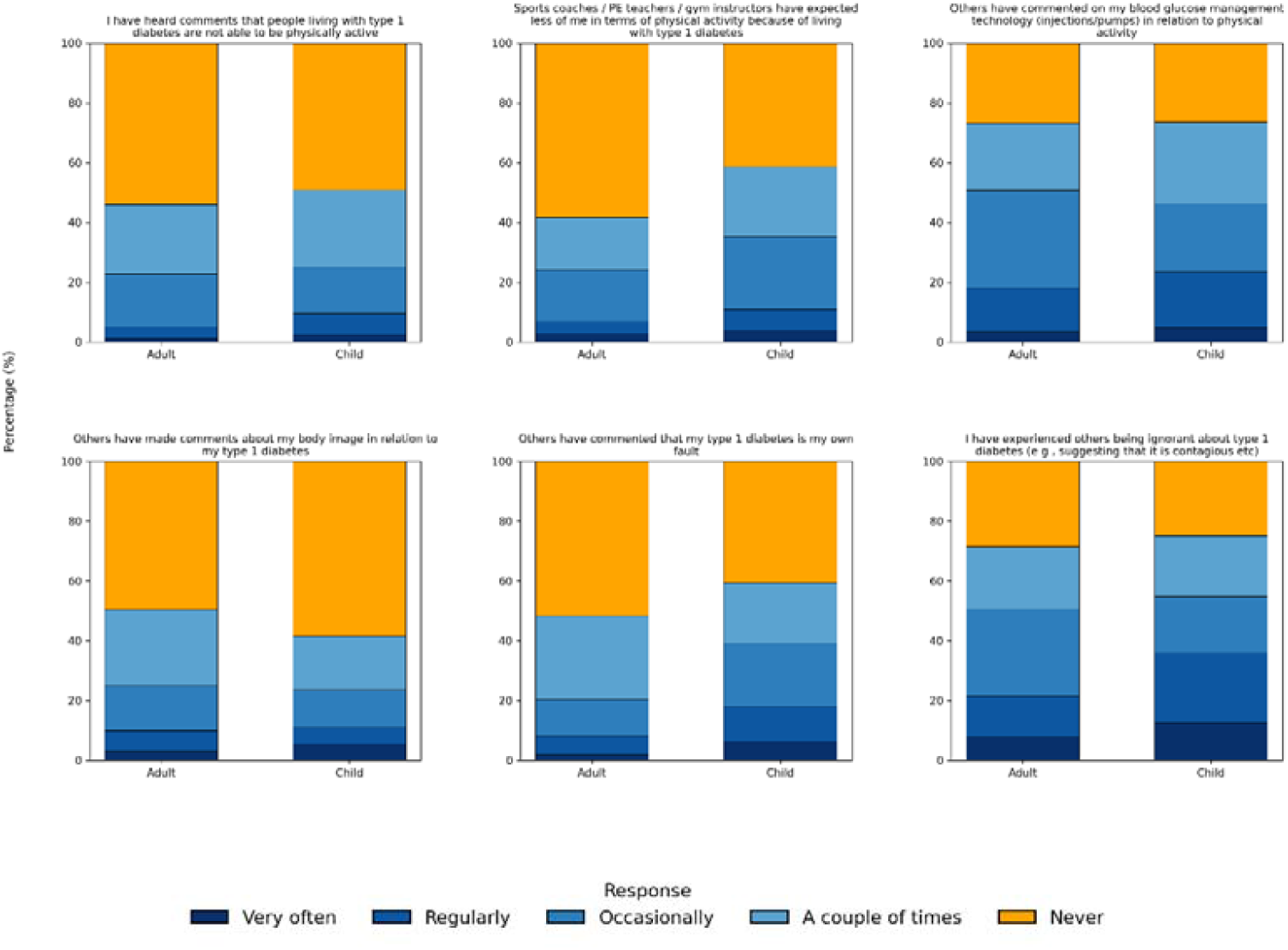
Participants experiences relating to perceived stigma and negative comments about T1D. Experiences related to physical activity and type 1 diabetes among adults and children. Stacked bar charts show the percentage of respondents (adults and parent-proxy responses for children) who reported each experience related to support or information about physical activity and type 1 diabetes. Responses are colour-coded by frequency

In relation to support from others, 42% of participants (48% of adults and 31% of children) reported that coaches, PE teachers, or gym instructors had never demonstrated any knowledge of type 1 diabetes in the context of PA. Only 37% of participants overall reported receiving regular or frequent support from their healthcare team regarding PA—this was notably higher among children (57%) compared to adults (24%).

Additionally, a significant proportion of respondents reported difficulty accessing relevant information: one-third of adults (33%) and nearly a quarter of children (23%) stated they were unable to find accessible guidance on PA in relation to type 1 diabetes. A more detailed breakdown of these findings is presented in Figure 5.

**Figure 5.**
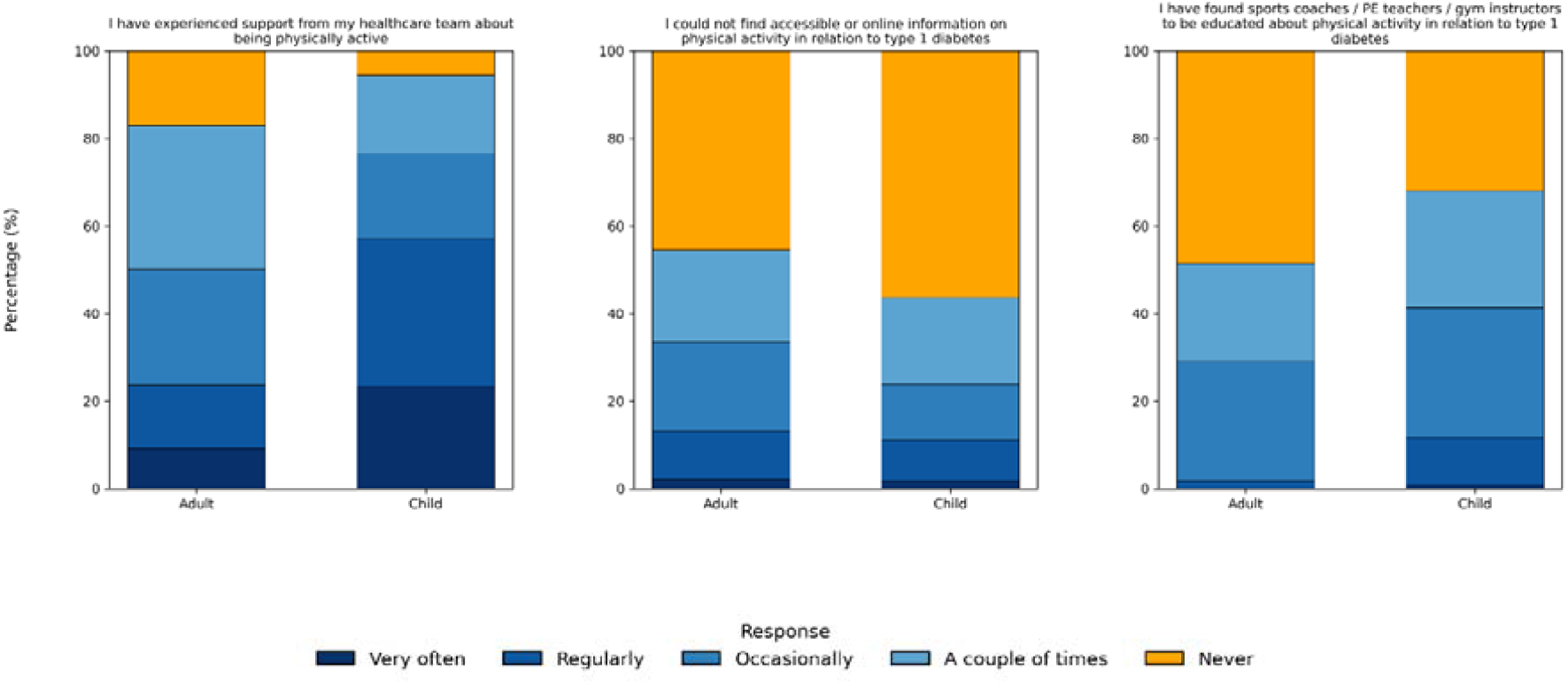
Participants experience of support and information accessibility. Experiences of stigma related to physical activity and type 1 diabetes, reported by adults and children. Stacked bar charts illustrate the percentage of participants (adults and parent-proxy responses for children) who reported experiencing various forms of stigma in relation to physical activity and type 1 diabetes. Responses are colour-coded by frequency

**Figure 6.**
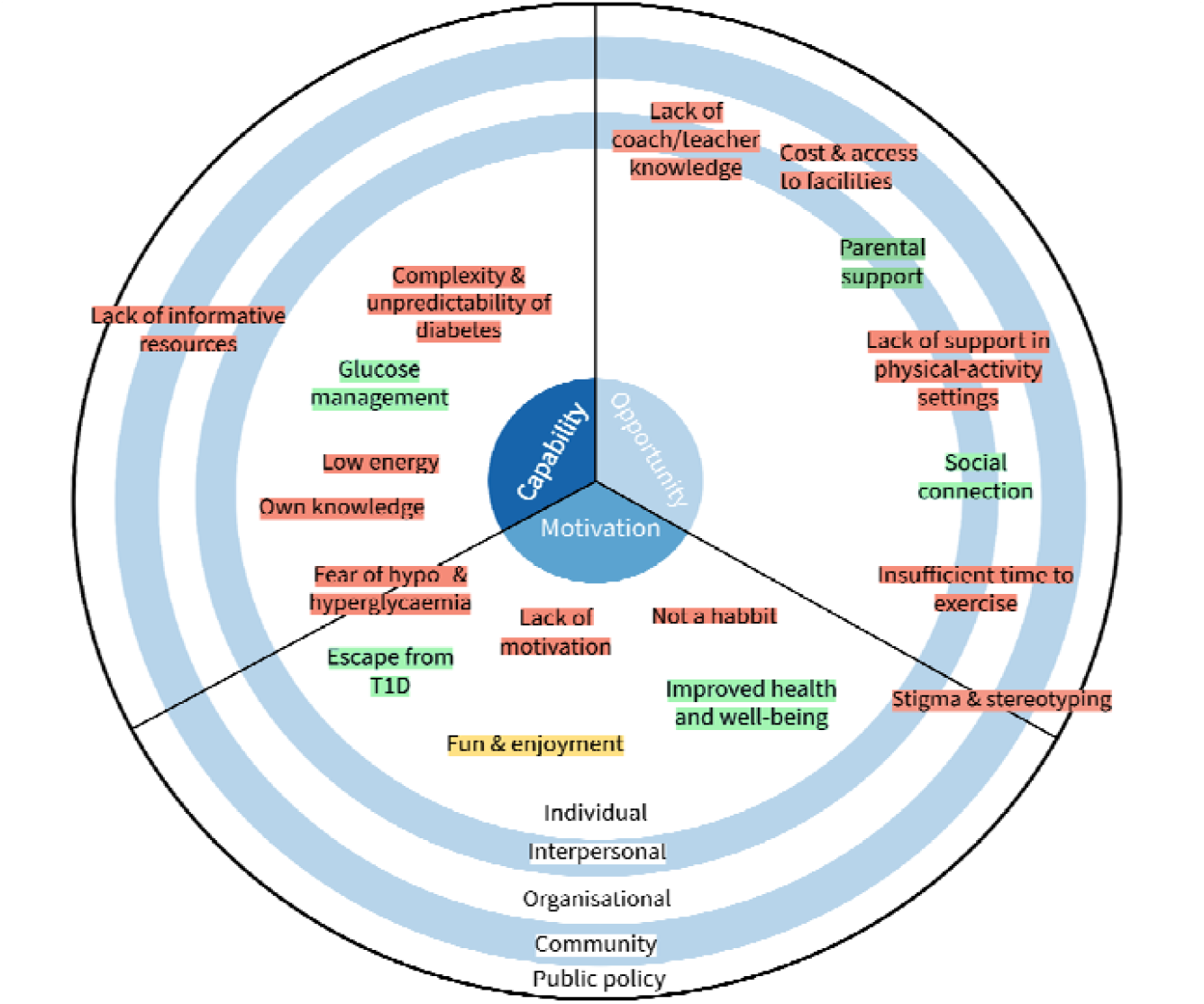
Summary of barriers and enablers to physical activity in individuals with T1D. Mapping of barriers and enablers to physical activity in people with type 1 diabetes using the COM-B model and socioecological framework. This figure presents the synthesised findings from a national survey of adults with type 1 diabetes and parents of children with T1D, displaying key barriers (red), enablers (green), and mixed factors (amber) influencing physical activity. Factors are positioned according to the COM-B behavioural model domains of Capability, Opportunity, and Motivation, and are further layered across the socioecological model levels: Individual, Interpersonal, Organisational, Community, and Public Policy. The positioning of each label reflects the dominant behavioural determinant and the structural level at which the factor operates.

### Qualitative Insights into factors influencing physical activity

Thematic analysis of free-text survey responses identified key barriers and motivators influencing PA in people with T1D.

#### Barriers to Physical Activity

Barriers were grouped into three main areas: diabetes-specific, interpersonal, and general. Diabetes-specific challenges were most frequently cited, with many participants pointing to blood glucose fluctuations and the unpredictability of diabetes as major obstacles. Fear of hypoglycaemia was a common concern *(“Having to stop activities when your blood sugar is too low*.*”)*, alongside frustration with glycaemic instability *(“It makes me very angry that they are so unreliable!”)*. The logistical burden of ongoing diabetes management, especially the need for constant planning, was seen to limit spontaneity (“Sometimes I’m unable to be spontaneous. Even with a pump, I still need to plan everything I do.”).

Social barriers often involved a lack of understanding from PA providers. Participants described being dismissed or misunderstood by coaches, teachers, and gym staff *(“Sometimes people see managing T1 diabetics as too much trouble and not worth the effort*.*”)*. Experiences of stigma and stereotyping were also reported: *“Comments about me being a recreational drug user because I’ve had to do injections in front of teammates*.*”*

Participants also reported common barriers such as lack of time, poor weather, low energy, and limited motivation, often exacerbated by the complexities of diabetes management.

#### Motivators for Physical Activity

Motivators for PA clustered into health and fitness, and psychological or social benefits. Many participants described improved physical and mental health as major drivers. The positive impact of activity on diabetes management was highlighted as a source of motivation *(“I can see the difference in my blood glucose management when I’m regularly active—it motivates me to keep going”)*. Others were motivated by body image, weight management, and maintaining general fitness.

Psychological and social factors also encouraged participation, including enjoyment (“Above all, I participate in my chosen sports because of enjoyment—it all puts a smile on my face”), social connection *(“It’s sociable and you meet like-minded people”)*, and personal challenge (“I enjoy a challenge and seeing reward for my training.”).

In summary, these findings illustrate the diverse and interconnected personal, social, and logistical factors shaping PA for people with T1D, with barriers and motivators often overlapping and context-specific.

### Synthesis of findings – future intervention suggestions

Table 2 and Figure 5 collectively present a synthesis of key barriers and enablers to PA in people with T1D, integrating findings from both the COM-B model and the socioecological model (SEM). These visual and tabulated summaries map behavioural influences across domains of capability, opportunity, and motivation, and across structural levels from the individual to the policy level. While some barriers relate to individual factors such as knowledge and confidence in managing blood glucose during exercise, many respondents described challenges that reflect broader systemic issues. These include limited awareness among coaches and teachers, experiences of stigma, and practical barriers such as cost and access—highlighting the need for multi-level, system-informed approaches.

**Table 2.**
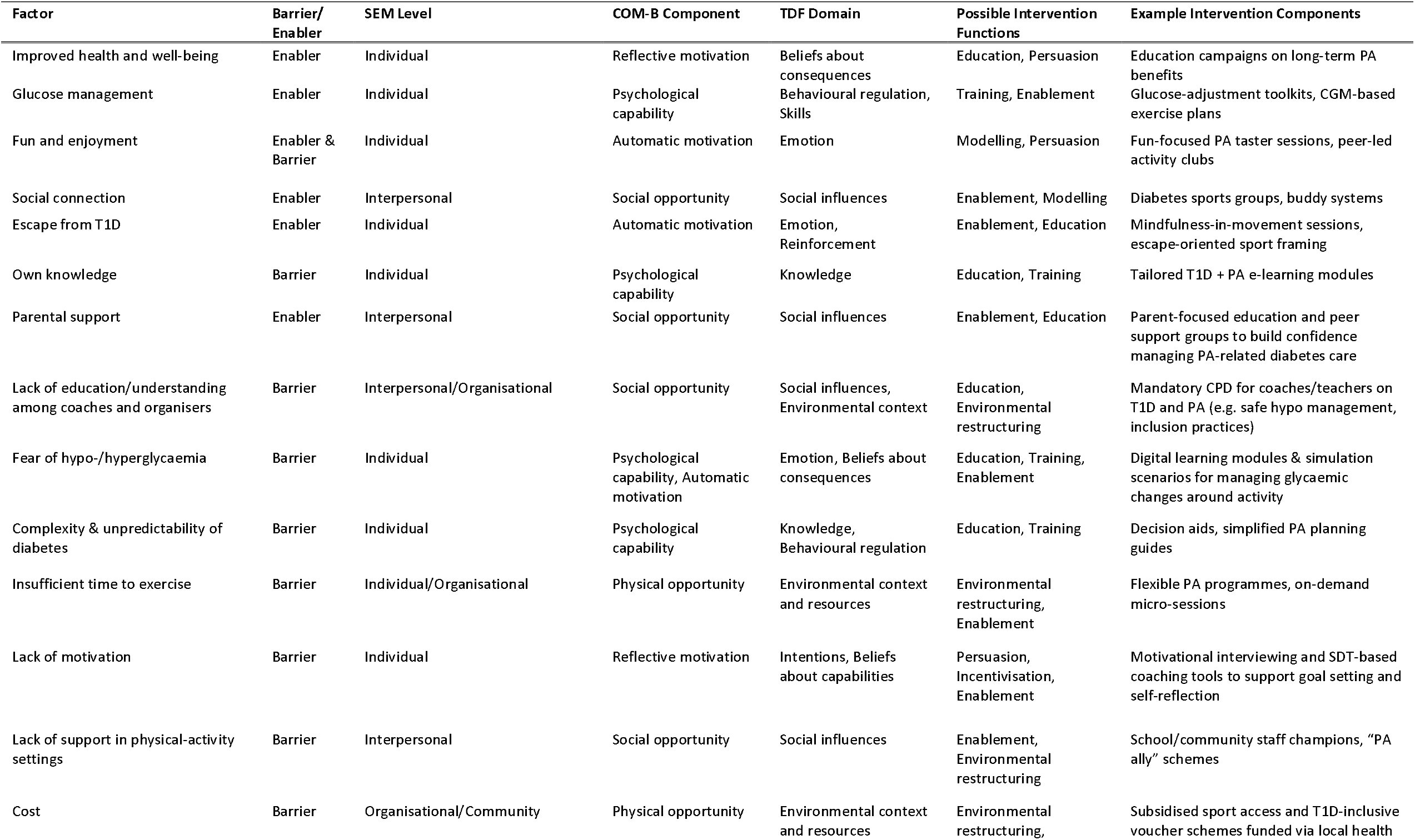
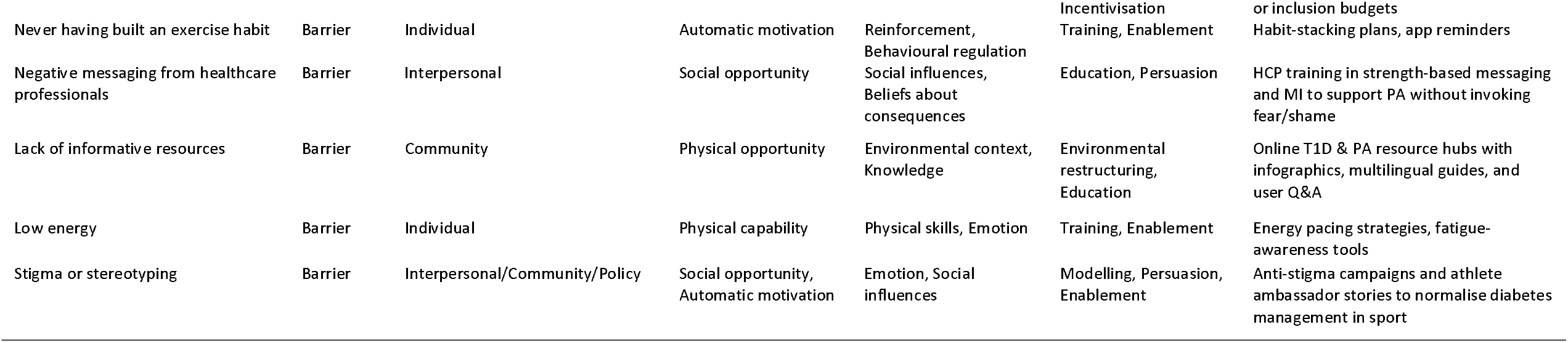
Mapping of barriers and enablers to physical activity in people with type 1 diabetes, aligned with socioecological and behavioural theory frameworks. Factors were identified through thematic analysis of survey data and mapped to the COM-B model and Theoretical Domains Framework (TDF) to inform intervention design. Intervention functions are drawn from the Behaviour Change Wheel (BCW), and example components reflect evidence-based or theory-informed strategies suitable for individual, community, and policy-level implementation. This synthesis highlights the need for multi-level, tailored approaches to support physical activity engagement in people with T1D.

Drawing on the Behaviour Change Wheel and Theoretical Domains Framework, several intervention strategies are suggested. These include: education and training to support both people with T1D and PA providers; enablement strategies such as personalised planning tools, peer mentoring, and parent or carer support networks; environmental restructuring to improve access and inclusivity in PA settings; and persuasive and modelling approaches to challenge stigma and promote more positive diabetes-related identities.

## DISCUSSION

This national survey is the first to directly compare barriers and enablers to PA among both adults and children living with type 1 diabetes (T1D), offering critical insights to guide future intervention design. While individual-level barriers—such as fear of hypoglycaemia and low motivation—remain significant, particularly in adults, the findings also highlight the powerful impact of external factors. Inadequate knowledge and support from coaches and educators, alongside pervasive experiences of stigma and stereotyping, emerged as key obstacles. These social-environmental barriers, often overlooked in previous research, featured prominently in participants’ accounts. The identification of stigma as a widespread barrier to PA is both novel and concerning, underscoring the urgent need for targeted education and systemic policy reform. To be effective, interventions must address not only individual behaviours but also the broader social and structural contexts in which people with T1D live and exercise.

Our findings in adults are consistent with those of previous qualitative research. Lascar et al. (2014), in a series of in-depth interviews with adults living with longstanding type 1 diabetes, identified six principal barriers to PA: lack of time and work-related factors, access to facilities, lack of motivation, embarrassment and body image concerns, weather, and diabetes-specific barriers such as limited knowledge about managing diabetes around exercise (18).

Our findings on barriers and facilitators to PA in children and adolescents with T1D closely reflect those of Dash et al (6). Their systematic review found that participation is influenced by individual characteristics, self-management demands, and the support of family, peers, teachers, and healthcare professionals. Similarly, our survey highlights that children are particularly affected by external barriers, such as limited support and understanding from coaches and teachers, alongside the practical challenges of blood glucose management during activity. This reinforces the importance of both individual and environmental factors in supporting PA for young people with T1D.

A novel and concerning finding of this study is the prominence of stigma and stereotyping as significant barriers to PA among individuals with type 1 diabetes While stigma—including blame, negative judgement, exclusion, and discrimination—has been described in broader T1D contexts (19). Our results demonstrate its direct and detrimental impact on PA engagement. Stigma has the potential to undermine social relationships, self-identity, emotional wellbeing, and diabetes self-management. It may also lead individuals to conceal their condition, thereby limiting participation in health-promoting behaviours. This is supported by systematic review evidence showing that stigma adversely affects self-care practices, uptake of diabetes technologies, and willingness to disclose one’s condition across both adult and paediatric population (20).

The high proportion of participants reporting negative comments or low expectations from coaches and teachers underscores the urgent need for education and awareness at interpersonal and community levels. Stigma related to type 1 diabetes (T1D) exerts a broad and pervasive impact, restricting access to education, employment, and opportunities for PA and participating in sports. Although the Equality Act 2010 provides legal protection against discrimination in certain sectors, our findings reveal a persistent gap between policy and lived experience within PA and sport, with many reporting ongoing discrimination and inadequate adjustments in schools, workplaces, and community settings (21). These results point to the need for enhanced advocacy efforts, including consideration for PA and sport to be explicitly addressed within anti-discrimination legislation.

In addition to stigma, lack of support and knowledge among coaches, PE teachers, and gym instructors remains a key barrier—an issue highlighted in both our current and previous research (22). This challenge appears particularly pronounced among children and adolescents, who rely on adults for decision-making and safety during PA. Adults, by contrast, may feel solely responsible for managing their diabetes, shaped by years of self-management or limited exposure to supportive environments. Sadly some participants were resigned to the lack of awareness among providers, perceiving it as an accepted norm. These variations in experience and expectations between age groups reinforce the need for targeted community-level interventions to enhance diabetes literacy among PA providers. Such initiatives are essential not only to improve practical competence but also to foster more inclusive, supportive, and equitable environments for individuals with T1D across the lifespan.

## STUDY LIMITATIONS

This study has several limitations. The use of a convenience sample recruited via a diabetes charity network may have introduced selection bias, with overrepresentation of individuals more engaged with diabetes organisations and a high proportion of technology users (20) limiting generalisability.

All data were self-reported and may be influenced by recall or social desirability bias. Parent-proxy responses for children may not fully reflect children’s own experiences, and qualitative analysis by a single researcher carries a risk of interpretive bias, though coding was discussed within the team.

Despite these limitations, the large national sample and mixed-methods design provide valuable insights into the multi-level factors influencing PA in people with T1D and inform priorities for future research and intervention development.

## RESEARCH/POLICY IMPLICATIONS

These findings highlight the need for multi-level, tailored interventions to support PA in people with T1D. For adults, strategies should focus on building confidence, addressing hypoglycaemia fears, and supporting self-management through education and personalised tools. For children and adolescents, the emphasis should be on creating supportive environments, with targeted training for coaches and teachers to reduce stigma and improve understanding.

Interventions should operate across individual, interpersonal, organisational, and community levels, supported by policy measures such as embedding diabetes education in schools and sports settings. Future research should prioritise co-designed approaches with people with T1D, families, and key stakeholders to ensure interventions are relevant, acceptable, and sustainable.

## CONCLUSION

Collectively, these findings underscore the urgent need for multi-level interventions to address stigma and stereotyping. Educational initiatives targeting the general public, healthcare professionals, educators, and sports coaches are essential to challenge misconceptions and promote understanding of T1D. Policy measures to enforce anti-discrimination legislation and ensure reasonable adjustments in all settings are also critical. Addressing stigma is not only a matter of individual wellbeing but a fundamental issue of equity and inclusion for people living with T1D.

## Data availability statement

This study involved secondary analysis of an existing dataset. Data available from third party upon reasonable request.

## Ethics statement

This study involved secondary analysis of an existing dataset. The study was conducted according to the ethical guidelines of the Declaration of Helsinki.

## Acknowledgments

Our sincere thanks to all participants who took part in the original survey. Acknowledgements Lead authors (EC) time was funded as part an National Institute for Health and Care Research (NIHR) School for Primary Care Research Postdoctoral Fellowship (C010)

For the purpose of open access, the author has applied a Creative Commons Attribution (CC BY) licence to any Author Accepted Manuscript version arising from this submission’

